# Towards Targeted Dementia Prevention: Population Attributable Fractions and Risk Profiles in Germany

**DOI:** 10.1101/2025.07.27.25332278

**Authors:** Iris Blotenberg, Jochen René Thyrian

**Affiliations:** Brigham and Women’s Hospital, Harvard Medical School, Boston, Massachusetts, USA; German Center for Neurodegenerative Diseases (DZNE), Greifswald, Germany; Institute for Community Medicine, Greifswald, Germany

**Keywords:** Alzheimer’s disease, cognition, cognitive decline, modifiable risk factors, lifestyle, prevention, risk groups

## Abstract

**INTRODUCTION:** Effective dementia prevention requires understanding the distribution of modifiable risk factors and identifying high-risk subgroups. We estimated the prevention potential in Germany and identified empirical risk profiles to inform precision public health.

**METHODS:** We analyzed nationally representative data from the 2023 German Ageing Survey. Population Attributable Fractions (PAFs) and Potential Impact Fractions (PIFs) were computed for established modifiable risk factors. Latent Class Analysis identified population-based risk profiles.

**RESULTS:** An estimated 36% of dementia cases in Germany are attributable to modifiable risk factors. Reducing their prevalence by 15–30% could prevent 170,000–330,000 cases by 2050. We identified four distinct risk profiles – metabolic, sensory impairment, alcohol consumption, and lower-risk – each associated with demographic and regional characteristics.

**DISCUSSION:** Our findings highlight considerable national prevention potential and reveal population subgroups with shared risk patterns. These profiles provide a novel foundation for designing targeted, equitable, and more efficient dementia prevention strategies.

## 1 BACKGROUND

Our understanding of modifiable dementia risk factors is rapidly evolving. The Lancet Commission’s updated report identified 14 modifiable risk factors across the lifespan, estimating that up to 45% of dementia cases worldwide may be linked to these factors [1]. The updated Lancet Commission report also presents revised estimates of relative risks, with higher values for depression and diabetes and lower ones for hearing loss, obesity, hypertension and smoking than in previous reports [2, 3].

In Germany, an estimated 1.8 million people are currently living with dementia [4, 5]. Without effective preventive measures, this number could rise to 2.7 million by 2050 [5]. In light of this, an up-to-date assessment of the national prevention potential is urgently needed. The release of new data from the 2023 wave of the German Ageing Survey (DEAS) – a nationally representative cohort – now enables such an assessment based on current prevalence rates and updated risk estimates [6]. The first aim of this study is to quantify the proportion of dementia cases in Germany attributable to modifiable risk factors and to estimate the potential impact of population-level risk reduction strategies.

While multidomain interventions, such as the FINGER trial and its international adaptations [7-10], have shown promise, such individual-level measures tend to have limited “population impact” due to narrow reach and the neglect of structural and environmental causes of disease [11, 12]. Although the assessment of the prevention potential at the national level is crucial, a universal or large-scale rollout of preventive strategies would be highly resource-intensive and risk misallocating efforts. Given the heterogeneity of dementia risk across the population [13], a more effective approach may lie in identifying and targeting empirically defined subgroups that share distinct constellations of modifiable risk factors.

There is a lack of studies using nationally representative data to systematically identify such dementia risk profiles in the population. The second aim of our study is therefore to derive data-driven risk subgroups using latent class analysis (LCA) and to examine their sociodemographic correlates. This approach can inform more precise, feasible, and equitable public health strategies and may provide a model for risk stratification in other countries.

## 2 METHODS

### 2.1 Risk factors and sociodemographic predictors

This study utilized data from the most recent wave (2023) of the German Ageing Survey (DEAS), published in March 2025 (n = 4,992) [6]. The DEAS is a nationwide representative cross-sectional and longitudinal survey of the German population aged 40 and older. Participants were interviewed either in person or by telephone using structured questionnaires and completed a written self-report. Of the 14 modifiable risk factors identified by the *Lancet Commission*, 12 were assessed; data on traumatic brain injury and air pollution were not available. Air pollution exposure could only be roughly approximated using district type. e*Table 1* in the Supplement provides an overview of the included risk factors. Sociodemographic characteristics included age, sex, district type, region (east / west) and cohabitation status.

**Table 1.** Prevalences, relative risks, commonalities, population attributable fractions (PAF), and potential impact fractions (PIF, for a 15% or 30% prevalence reduction) for twelve potentially modifiable risk factors.

| Risk factor | Prevalence in the population in Germany, % | Relative risk [95% CI] | Communality, % | PAF, % [95% CI] | Adjusted PAF, % [95% CI] | 15% reduction of risk factor prevalence |  | 30% reduction of risk factor prevalence |  |
| --- | --- | --- | --- | --- | --- | --- | --- | --- | --- |
|  |  |  |  |  |  | PIF, % [95% CI] | Adjusted PIF, % (95% CI) | PIF, % [95% CI] | Adjusted PIF, % (95% CI) |
| Less education | 15.4 | 1.6 (1.3-2.0) | 46.2 | 8.5 (4.4-13.3) | 3.3 (2.0-4.5) | 1.3 (0.7-2.0) | 0.6 (0.3-0.9) | 2.5 (1.3-4.0) | 1.1 (0.6-1.7) |
| Hearing loss | 23.3<sup>1</sup> - 40.6<sup>2</sup> | 1.4 (1.0-1.9) | 56.7 | 14.0 (0.0-26.8) | 5.4 (0.0-9.1) | 2.1 (0.0-4.0) | 0.9 (0.0-1.8) | 4.2 (0.0-8.0) | 1.8 (0.0-3.4) |
| High LDL cholesterol | 23.9 | 1.3 (1.3-1.4) | 43.5 | 6.7 (6.7-8.7) | 2.6 (3.0-3.0) | 1.0 (1.0-1.3) | 0.5 (0.5-0.6) | 2.0 (2.0-2.6) | 0.9 (1.0-1.1) |
| Depression | 10.3<sup>3</sup> – 15.9<sup>4</sup> | 2.2 (1.7-3.0) | 59.2 | 16.0 (10.0-24.1) | 6.2 (4.5-8.2) | 2.4 (1.5-3.6) | 1.1 (0.7-1.6) | 4.8 (3.0-7.2) | 2.1 (1.4-3.0) |
| Physical inactivity | 20.7 | 1.2 (1.2-1.3) | 33.3 | 4.0 (4.0-5.8) | 1.5 (1.8-2.0) | 0.6 (0.6-0.9) | 0.3 (0.3-0.4) | 1.2 (1.2-1.8) | 0.5 (0.6-1.0) |
| Smoking | 22.3 | 1.3 (1.2-1.4) | 55.3 | 6.3 (4.3-8.2) | 2.4 (1.9-2.8) | 0.9 (0.6-1.2) | 0.4 (0.3-0.5) | 1.9 (1.3-2.5) | 0.8 (0.6-1.0) |
| Diabetes | 11.1 | 1.7 (1.6-1.8) | 51.3 | 7.2 (6.2-8.2) | 2.8 (2.8-2.8) | 1.1 (0.9-1.2) | 0.5 (0.5-0.5) | 2.2 (1.9-2.4) | 1.0 (0.9-1.0) |
| Hypertension | 34.5 | 1.2 (1.1-1.4) | 60.0 | 6.5 (3.3-12.1) | 2.5 (1.5-4.1) | 1.0 (0.5-1.8) | 0.4 (0.2-0.8) | 1.9 (1.0-3.6) | 0.9 (0.5-1.5) |
| Obesity | 26.7 | 1.3 (1.0-1.7) | 59.5 | 7.4 (0.0-15.7) | 2.9 (0.0-5.4) | 1.1 (0.0-2.4) | 0.5 (0.0-1.0) | 2.2 (0.0-4.7) | 1.0 (0.0-2.0) |
| Alcohol consumption | 29.2 | 1.2 (1.0-1.5) | 60.7 | 5.5 (0.0-12.7) | 2.1 (0.0-4.3) | 0.8 (0.0-1.9) | 0.4 (0.0-0.8) | 1.7 (0.0-3.8) | 0.7 (0.0-1.6) |
| Social isolation | 8.9 | 1.6 (1.3-1.8) | 31.8 | 5.1 (2.6-6.6) | 2.0 (1.2-2.3) | 0.8 (0.4-1.0) | 0.3 (0.2-0.4) | 1.5 (0.8-2.0) | 0.7 (0.4-0.8) |
| Visual impairment | 12.1 | 1.5 (1.4-1.6) | 62.0 | 5.7 (4.6-6.8) | 2.2 (2.1-2.3) | 0.9 (0.7-1.0) | 0.4 (0.3-0.4) | 1.7 (1.4-2.0) | 0.8 (0.7-0.8) |
| Total |  |  |  |  | **35.7**<br>**(20.6-50.8)** |  | **6.3**<br>**(3.4-9.8)** |  | **12.2**<br>**(6.6-18.7)** |

*Notes.* Commonality represents the proportion of shared variance of the risk factor in question and the other risk factors. <sup>\*1</sup> self-reported hearing impairment from the German Aging Survey (n = 4992) <sup>\*2</sup> Prevalence from Döge (1) <sup>\*3</sup> Point prevalence from the German Aging Survey (2), <sup>\*4</sup> Lifetime prevalence from the NAKO Health Study (3); CI, confidence interval

### 2.2 Statistical analysis

#### Prevention Potential

Prevention potential was estimated following the Lancet Commission’s approach [1, 3], using adjusted Population Attributable Fractions (PAFs) via a modified Levin’s formula [14]. Relative risks were drawn from the most recent Lancet report [1]; prevalence estimates from DEAS 2023 [6]. Post-stratified weights based on the 2023 Microcensus were applied to ensure representativeness. Weighting factors adjusted for risk factor intercorrelations (e.g., metabolic syndrome components). A principal component analysis (PCA) of the tetrachoric correlation matrix was used to derive the weights, defined as one minus the communality for each risk factor. Potential Impact Fractions (PIFs) were also calculated to estimate case reductions under hypothetical risk factor reductions of 15% and 30%.

#### Identification and characterization of risk profiles

Profiles of risk factors were identified using LCA. This statistical method, based on a structural equation modeling framework, allows for the identification of homogeneous subgroups within a heterogeneous sample. We applied full information maximum likelihood (FIML) to handle missing data in the risk factor variables. The highest proportion of missing data was observed for the variable hearing loss, with 932 missing values (18.7%). We tested models with one to five classes. A total of eleven dementia-related risk factors – including all health and lifestyle factors – were used to define the LCA. The risk factor low education was included as a covariate, along with age, sex, living situation, district type and region (eastern or western Germany). A multinomial logistic regression was performed to examine the associations between these covariates and the latent risk profiles, with the aim of identifying public health-relevant subgroups. Model selection was based on the Akaike Information Criterion (AIC), Bayesian Information Criterion (BIC), sample-size adjusted BIC (ssABIC), Vuong-Lo-Mendell-Rubin likelihood ratio test (VLMRT) and the Lo-Mendell-Rubin adjusted likelihood ratio test (LMRT), as well as interpretability. All analyses were conducted using Mplus version 8.11 [15].

## 3 RESULTS

### 3.1 Prevention Potential

*Table 1* shows the prevalences, relative risiks, communalities, PAF und PIF for 12 of the 14 potentially modifiable risk factors for dementia identified by the Lancet Commission. Overall, the potential for dementia prevention in Germany was estimated at 36%. When including the risk factor air pollution, which could only be approximated using the district type, the prevention potential increased to 39% (see Supplement *eTable2*). The most influential risk factors were depression, hearing impairment, low educational attainment, overweight and diabetes.

*Figure 1* illustrates the projected number of dementia cases that could be prevented by 2050, if the population-level prevalence of these risk factors were reduced by 15% or even 30%. A 15% reduction could lead to 170,000 fewer dementia cases by 2050 (from the projected 2.7 million cases), while a 30% reduction could potentially prevent up to 330,000 cases.

**Figure 1.**
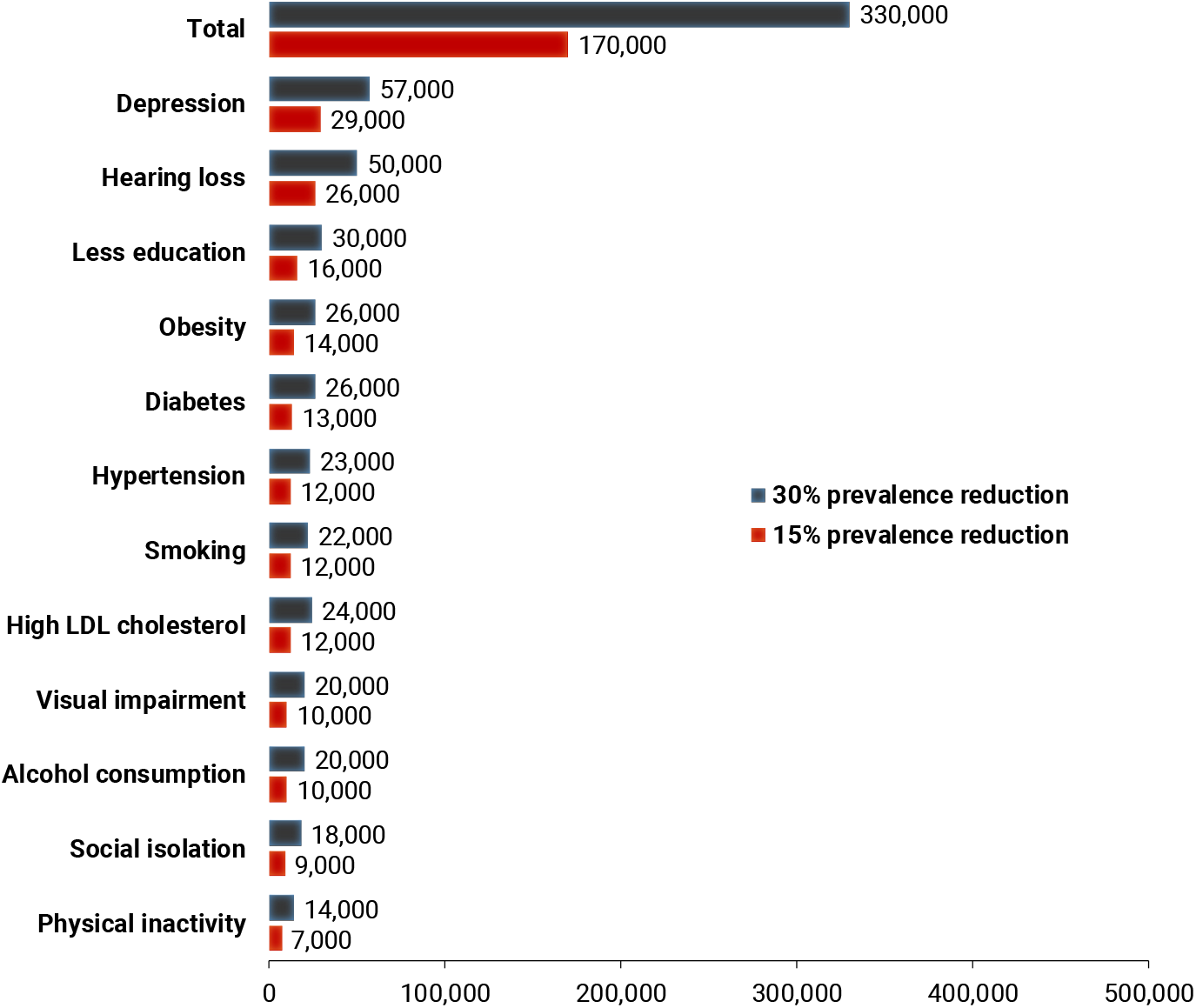
Number of dementia cases in 2050 that could theoretically be prevented if the population-based prevalence of twelve modifiable risk factors were to be reduced by 15% or 30%. Basis for calculation: 2.7 million people with dementia in 2050. [please use color in print]

### 3.2 Risk profiles

Based on model fit indices, the four-class model was selected as the best-fitting solution (see Supplement *eTable3* for the goodness-of-fit indices). The risk profiles of these four classes are presented in *Figure 2*. They were labeled according to the distribution of risk factors as follows: 1) The “metabolic syndrome” profile was characterized by the highest probabilities of hypertension, overweight, elevated LDL cholesterol and diabetes. 2) The “sensory impairment” profile showed the highest probabilities for hearing loss and visual impairment. 3) The “alcohol consumption” profile had the highest probability of alcohol use. 4) The “lower risk” profile was characterized by probabilities below 0.3 for all risk factors.

**Figure 2.**
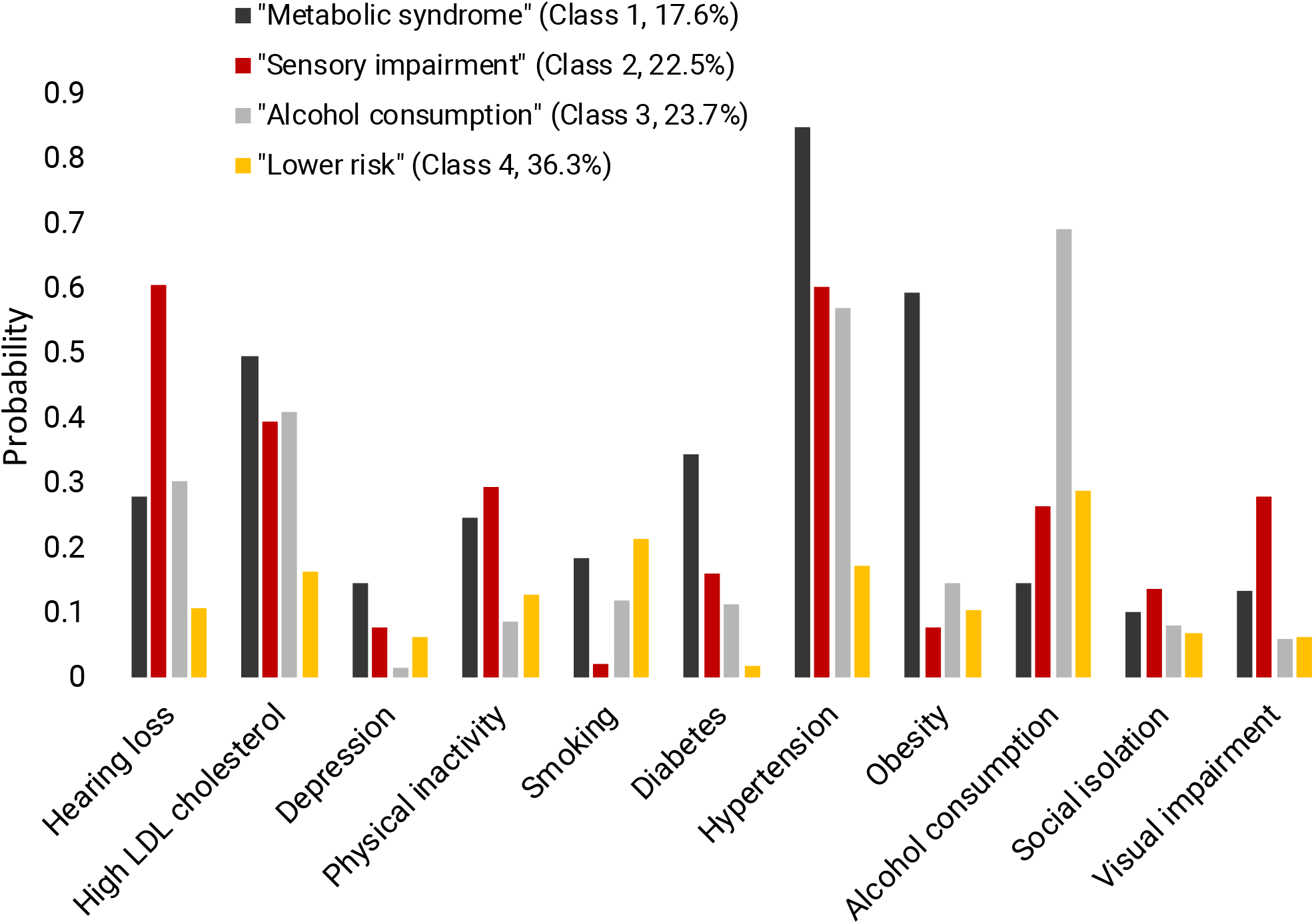
Probabilities of the presence of risk factors for each risk factor profile. [please use color in print]

#### Characterization of risk profiles

The results of the multinomial logistic regression predicting risk profile membership are presented in *Table 2*. Individuals in the “metabolic syndrome” risk profile were older, had lower educational attainment and were more than twice as likely to live in eastern Germany compared to individuals in the “lower risk” profile. They were also more likely to live in smaller towns or rural districts. Individuals in the “sensory impairment” profile were also older, lived more frequently in eastern Germany and were more likely to live in smaller towns than those in the “lower risk” profile. Finally, individuals in the “alcohol consumption” profile were older, more often male and more likely to be cohabiting compared to those in the “lower risk” group. *eTable 4* in the Supplement shows further details on the sociodemographic characteristics of the four risk profiles.

**Table 2.** Relative risks and 95% confidence intervals from the latent class analysis model with covariates using the lower risk group (Class 4) as the reference (n = 4989)

| Variable | Class 1 „Metabolic syndrome“ |  |  |  | Class 2 „Sensory impairment“ |  |  |  | Class 3 „Alcohol consumption“ |  |  |  |
| --- | --- | --- | --- | --- | --- | --- | --- | --- | --- | --- | --- | --- |
|  | RR | 95% CI |  | *p* | RR | 95% CI |  | *p* | RR | 95% CI |  | *p* |
| Age | 1.082 | 1.041 | 1.125 | **<0.001** | 1.363 | 1.272 | 1.461 | **<.001** | 1.126 | 1.077 | 1.177 | **<.001** |
| Sex |  |  |  |  |  |  |  |  |  |  |  |  |
| Female | 0.559 | 0.202 | 1.546 | 0.262 | 0.480 | 0.172 | 1.343 | 0.162 | 0.170 | 0.073 | 0.396 | **<.001** |
| Education level |  |  |  |  |  |  |  |  |  |  |  |  |
| Moderate education | *0.517* | *0.266* | *1.008* | **0.053** | 1.807 | 0.588 | 5.551 | 0.302 | 0.872 | 0.299 | 2.544 | 0.803 |
| High education | 0.227 | 0.078 | 0.657 | **0.006** | 1.924 | 0.527 | 7.019 | 0.322 | 1.818 | 0.477 | 6.928 | 0.381 |
| Living situation |  |  |  |  |  |  |  |  |  |  |  |  |
| Cohabiting | 0.887 | 0.516 | 1.526 | 0.665 | 1.093 | 0.593 | 2.015 | 0.775 | 3.042 | 1.739 | 5.322 | **<.001** |
| Region |  |  |  |  |  |  |  |  |  |  |  |  |
| Eastern Germany | 2.313 | 1.594 | 3.356 | **<0.001** | 2.089 | 1.095 | 3.985 | **0.025** | 1.043 | 0.397 | 2.739 | 0.932 |
| District type |  |  |  |  |  |  |  |  |  |  |  |  |
| Independent large city (ref.) |  |  |  |  |  |  |  |  |  |  |  |  |
| Urban district | 1.507 | 1.044 | 2.175 | **0.029** | 1.842 | 1.003 | 3.383 | **0.049** | 1.483 | 0.683 | 3.216 | 0.319 |
| Rural district with densification trends | 1.241 | 0.826 | 1.865 | 0.299 | 1.524 | 0.714 | 3.252 | 0.276 | 1.062 | 0.432 | 2.613 | 0.895 |
| Sparsely populated rural district | 1.684 | 1.088 | 2.606 | **0.019** | 1.004 | 0.439 | 2.296 | 0.993 | 0.971 | 0.475 | 1.986 | 0.935 |

Notes. RR, relative risk; CI, confidence interval

## 4 DISCUSSION

In this nationally representative study, we show that there is a considerable prevention potential for dementia in Germany: an estimated 36% of dementia cases are attributable to 12 of the 14 modifiable risk factors identified by the Lancet Commission [1]. This estimate is lower than the global figure of 45% reported by the Lancet Commission, which can be attributed to differences in the distribution of risk factors. It is consistent with findings from other country- or region-specific analyses [e.g. 16, 17] and underscores the relevance of tailoring prevention strategies to specific population contexts. Notably, if a 15% reduction in risk factor prevalence – as targeted by the U.S. National Alzheimer’s Project Act (NAPA) – were achieved in Germany, approximately 170,000 dementia cases could be prevented or delayed by 2050. A 30% reduction could yield more than 330,000 prevented cases. Moreover, such measures may have broader effects on other conditions that share similar risk factors, such as heart disease, stroke, or cancer [18].

Beyond the overall prevention potential, we identified four distinct population-based risk profiles using LCA. These profiles – metabolic, sensory impairment, alcohol consumption, and lower risk – represent empirically derived constellations of modifiable dementia risk factors. To our knowledge, this is one of the first studies to identify such subgroups using nationally representative data. Importantly, the risk profiles were associated with sociodemographic characteristics such as age, education, and region, pointing toward structural differences in dementia risk exposure across the population. These findings lay a foundation for developing more targeted, subgroup-specific prevention strategies that go beyond a universal, one-size-fits-all model.

Several modifiable risk factors emerged as particularly influential: depression, hearing loss, low educational attainment, obesity, and diabetes. Many of these factors are influenced not only by individual behavior, but also by structural determinants of health. Effective dementia prevention in Germany will therefore require a combination of individual-level behavioral interventions and broader system-level strategies, such as improved access to mental health care, increased availability of hearing aids, and policies that reduce educational inequality. Education deserves special attention – not only as an independent protective factor, but also as a determinant of many other risk factors [e.g. 19]. In Germany, educational attainment is strongly associated with socioeconomic disparities [20]. Enhancing educational equity may therefore serve as an important lever for dementia prevention.

Our findings indicate structurally rooted regional differences in dementia risk. For instance, individuals living in rural areas and Eastern Germany were more likely to belong to high-risk profiles, particularly the metabolic or sensory impairment risk profiles. These results are also consistent with the regional distribution of dementia cases in Germany, where – even after age standardization – a particularly high prevalence is found in the eastern federal states [21], which tend to be structurally disadvantaged [22]. This knowledge of diverse dementia risk profiles and their sociodemographic patterns can inform more targeted and effective prevention strategies. For example, it highlights the need to intensify prevention efforts in rural areas and socioeconomically disadvantaged regions, with a particular focus on individuals with lower levels of education.

The identification of latent risk profiles in our study is consistent with findings from analyses using UK Biobank data [23]. There, similar profiles – particularly those related to metabolic health and substance use – have been identified. The convergence of findings across datasets and contexts suggests that these risk constellations may reflect broader population-level patterns. Future studies should explore the generalizability of these profiles across countries and cultural settings and test the feasibility and effectiveness of tailored intervention strategies for each subgroup.

### 4.1 Limitations

When calculating the prevention potential, methodological limitations must be considered: In its original form, Levin’s formula determines the prevention potential in isolation for a single risk factor [24]. In this analysis, we used a modified version with weighting factors to adjust for intercorrelations between risk factors [14]. However, potential interactions between risk factors (including non-modifiable ones) are not accounted for in the formula – this remains an area in need of further research [25]. A second limitation concerns selection bias – although the aim of the DEAS is to achieve a representative sample of the population, certain groups tend to participate disproportionately often in surveys (e.g., individuals with higher educational attainment) [26]. In the DEAS as well, individuals with a low level of education are underrepresented. To correct for selection bias and still estimate the prevalence of dementia risk factors as accurately as possible, weights based on the German Microcensus were applied.

## 5 CONCLUSION

Our analyses indicate that there is a substantial potential for dementia prevention in Germany and that a considerable number of cases could be prevented or delayed through effective interventions. In addition, we identified distinct risk profiles within the population, each defined by specific constellations of modifiable risk factors and sociodemographic characteristics. These profiles offer a valuable foundation for developing more targeted and equitable prevention strategies that are tailored to the needs of different population subgroups. Future research should focus on testing the effectiveness and feasibility of targeted interventions for empirically identified subgroups, taking both risk exposure and access barriers into account.

## Data Availability

The data used in this study are from the German Ageing Survey (DEAS), which is administered by the German Centre of Gerontology (DZA). The DEAS data are publicly available for scientific use upon application through the Research Data Centre of the DZA (FDZ-DZA). More information and access instructions can be found at: https://www.dza.de/en/fdz

## List of abbreviations

DEAS: German Ageing Survey
PAF: Potential Attributable Fraction
PIF: Population Impact Fraction
PCA: Principal component analysis
FINGER: Finnish Geriatric Intervention Study to Prevent Cognitive Impairment and Disability
LCA: Latent class analysis
FIML: Full information maximum likelihood
AIC: Akaike Information Criterion
BIC: Bayesian Information Criterion
VLMRT: Vuong-Lo-Mendell-Rubin likelihood ratio test
LMRT: Lo-Mendell-Rubin adjusted likelihood ratio test

## Acknowledgments

The authors received no additional support beyond their own contributions.

## Consent Statement

We used anonymized secondary data from the German Ageing Survey (DEAS). All participants provided written informed consent at the time of data collection. The study procedures complied with applicable ethical standards and data protection regulations.

